# The Pandemics in Artificial Society Agent-Based Model to Reflect Strategies on COVID-19

**DOI:** 10.1101/2020.07.27.20162511

**Authors:** Hokky Situngkir, Andhika Bernad Lumbantobing

## Abstract

Various social policies and strategies have been deliberated and used within many countries to handle the COVID-19 pandemic. Some of those basic ideas are strongly related to the understanding of human social interactions and the nature of disease transmission and spread. In this paper, we present an agent-based approach to model epidemiological phenomena as well as the interventions upon it. We elaborate on micro-social structures such as social-psychological factors and distributed ruling behaviors to grow an artificial society where the interactions among agents may exhibit the spreading of the virus. Capturing policies and strategies during the pandemic, four types of intervention are also applied in society. Emerged macro-properties of epidemics are delivered from sets of simulations, lead to comparisons between each policy/strategy,s effectivity.

## 1 Introduction

When it comes to coronavirus disease (COVID-19) pandemic, the world witnesses heterogeneous ways of managing. Since the beginning of the outbreak in many places, different ways of managing the virus spreadings reflect the various healthcare situations faced by each government in the world, due to the testing and hospitalizations facilities. The situations are getting more complex for the unique characteristics of the COVID-19 itself.

As illustrated in figure 1, the fatality rate and the contagiousness of the COVID-19 are in between many other known diseases. The fact that the infected persons have some degree of possibilities to be asymptomatic, as well as the recognized pre-symptomatic cases, made a direct impact on the social life on which the epidemic occurs. Most infected people have mild symptoms and still be able to deliver their daily social activities in which they can easily spreading the virus rapidly in the population. Thus, policies of those limiting social interactions for the whole population are the most suggested solutions epidemiologically, as well as closing borders between places (countries, provinces, districts, etc.) to avoid imported/exported cases between regions. The consequences of the policies would apparently give a direct impact on many social aspects since interactions are fundamental elements in social life (Smelser, 1997). Modern human life is formed in the patterns of social interactions, in which all social aspects are embedded, including religions, customs, economy, politics, and all.

**Figure 1.**
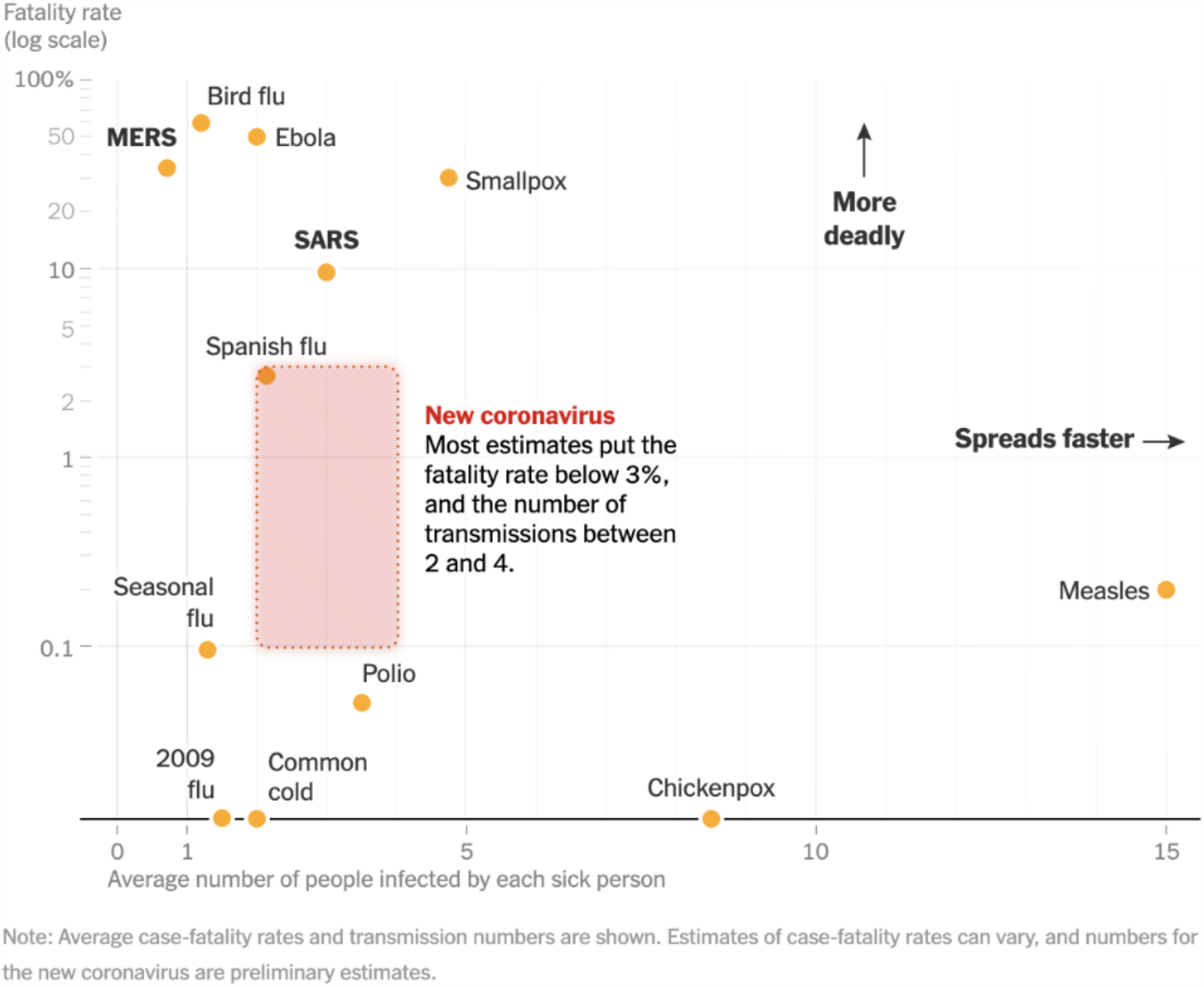
The comparisons of novel coronavirus to other epidemic diseases^1^.

Thus, the epidemiological suggestions for the policies, pro-social limitations and closing administrative borders are being resisted by the need of economical aspects of the social life. People are needed to be supported economically while on the other hand should be stay put with distant social interactions for the sake of damping the easily doubling trends of the infections. One popular policy is the “lock-down” within areas, where people are encouraged (and restricted) to “stay at home” (WHO, 2020). In contrast, some other countries enforce other kinds of the policy of not applying the “lock-down” at all, like in Sweden (Dahlber, *et. al*., 2020) with very mild restrictions due to social activities within the country. Many aspects happened to be put into account when it comes to social restrictions, from the reasons related to human rights to the economic aspects due to processes within the economy, macroeconomic reserve and workforces, and a lot more.

Furthermore, the considerations backing the varsities of enforced policies and strategies facing the COVID-19 may also be coming from the heterogeneous natural aspects of countries around the world. For instance, the interactive effects based on meteorological influences in the COVID-19 transmission and spread, due to the interactive effect between daily temperature and relative humidity on COVID-19 incidence (Qi, *et. al*., 2020). Some countries in tropical areas may have a milder effect within their infection cases relative to those in sub-tropical regions. The urge for maximum social restrictions in warmer tropical areas may not as great as those in other colder places. The exhibited variations are there for different existing situations faced countries in battling the pandemics.

The paper is preparing the toy model reflecting the social structure that in return presenting a proposal for enriching observations toward various strategies within many places due to the pandemic of COVID-19. While other previous works may present the computational models and simulations to approach the dynamics of an epidemic outbreak (*cf*. Situngkir, 2004), the coverage of the paper is focused on the preparations of artificial societies in which some policies may be grown (Epstein & Axtell, 1996) and explained. The model used tried to capture the micro-social structure on which the infected people came along. The people are interacting with one another with particular individual motives and move around the artificial world as lattices and grids (*cf*. Rhodes & Anderson, 1996). As an infected agent come along, the interactions exhibit the spreading of the virus in a sort of social network based on their bounded situations (*cf*. Newman, 2002). The model runs as an agent-based model (Gilbert & Terna, 2000) living the landscape that we can use to monitor the macro-properties of the epidemics (*cf*. Situngkir, 2003).

The observations are delivered in the emerged aspects of epidemics, *i*.*e*.: the number of people infected and how it spreads over the landscape as the complex adaptive system (Miller, 2007). If the behavior of the agents is related to the micro-level of description, the government policies can be seen as in the meso-one (medium level, between micro and macro). All of those aspects are then depicting the relations of “factors to actors” relations (Macy & Willer, 2002). Some possible conjectures in the ways to verify the results (Situngkir, 2004) are also discussed. Eventually, the paper is demonstrated how interdisciplinary works can enhance social policies handling pandemics (Angulo, 1987).

## 2 The Micro-social structure

We can see the micro-social used in the simulations as three parts, *i*.*e*.: the internal state of the agents, the mobility in our artificial world, and the spreading of the disease based on the first two properties.

### 1. Agent internal state

Agent *i* has internal state ruled by the social-psych-wellness index, denoted as *g*_*i*_(*t*) ∈ ℜ, *g*_*i*_(*t*) = [0.1]. This index represents the wellness of agents regarding many complex aspects in social life and it is composed of three factors, two of them are related directly to its corresponding spatial situations. The first one is denoted as *e*_*i*_(*t, x, y*), how agent *i* fulfill her necessities when she nears a static point, we can say this as attraction points located in 2-dimensional *x, y*. One can imagine these points as public surroundings where people meet and have their needs from others, *e*.*g*.: marketplace, recreational spaces, offices. Index *e*_*i*_(*t, x, y*) is determined by the euclidean distance to the nearest attraction point, adjusted by parameter *β*_*attraction*_. However, the relation is inversely proportional so that the nearer the agent to her chosen point, the higher *e*_*i*_(*t, x, y*). Its value is *e*_*i*_(*t, x, y*) ∈ ℜ = [0,1] by a negative exponential function, decaying with term 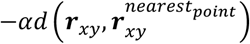.

The second aspect is the agent *i* neighborhood *n*_*i*_(*t, m*) which is simply a fraction of the number of her neighbors *m* divided by a global constant maximum neighbors M. The factor corresponds to understanding that human tends to reach out others for their individual needs, be it economic, cultural, and social well-being. The conceptualizations of the neighborhood are other agents as perceived within radii *d*_*neighbors*_, compared to her distance to others. This *n*_*i*_(*t, m*) ∈ ℜ = [0.1], weighted by a parameter *β*_*neighboring*_.

As a feedback of the growing index of social-psycho-wellness, there is a boredom factor with proportions to agent *i*,s, −*γg*_*i*_ (*t*). The higher the value of her gladness, the more this factor reducing *g*_*i*_(*t*). The three factors which determine the agent,s internal state can formally be stated by the following system of equations:

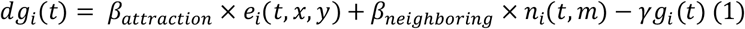

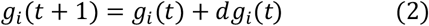

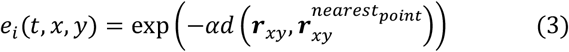

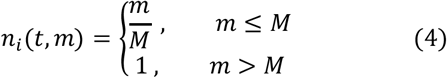

### 2. Agent spatial mobility

Our artificial world is 2-dimensional lattice and grids of which form torus in 3 dimensionalities since the edge of the left is glued with the right one, and the top with the bottom edge as well. Each agent spatial states are represented as a vector of position 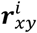 and her velocity 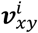. Initially, agents are randomly located uniformly with zero speed of movement. Following the terms of classical mechanics, in every iteration the spatial states of agents are updated by applying an amount of normalized steering force to agent *i*:

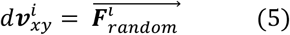

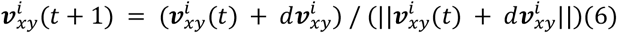

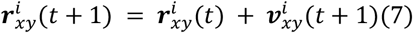

whereby default, each value of 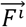 components is taken from an unbiased uniform distribution and the mass is assumed to be unity. Furthermore, some conditional force is computationally adapted based upon the agent *i* internal state, *g*_*i*_(*t*). Thus, the agent,s mobility is ruled by her micro-motives: maximizing the wellness index. In short, the dynamics adopt Boids Model (Reynold, 1987) which follows some simple rules distributed among the agents, the separation and cohesion-like rule.

Only if *g*_*i*_(*t*) is below the threshold value *g*_*low*_, we replace 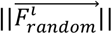 to 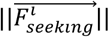 This force will make agents move towards their respective neighbors, average position and their corresponding nearest attraction points. However, dynamically, the neighboring factor is set to be zero when her number of neighbors m exceeds maximum limit *M* forbidding the overcrowded localities. This also happened when an agent has no neighboring agents. In case no attraction point that they can perceive, or no neighboring force, they will perform a random walk by default.

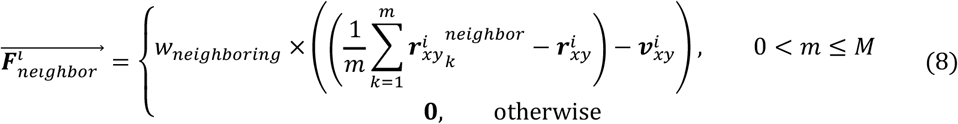

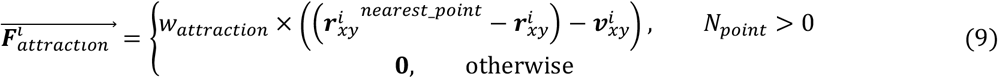

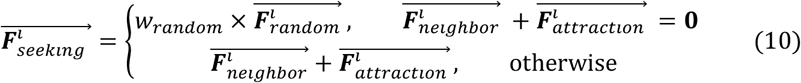

where *w* denotes the value of weighting factor times normalization factor for each force. Another rule is the separation rule which is necessary for later when we do the simulations. This rule makes agent *i* keep their distance from each other by applying force:

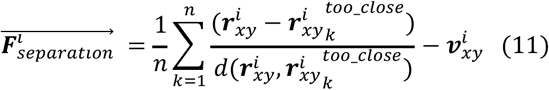

The equation corresponds to *d*_*separation*_value which determines how an agent takes into account their neighbors perceived as too close,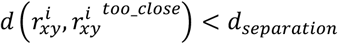. The denominator gives proportionality to the applied force so that the nearer her neighbor, the stronger she will stay away. To summarize, the change of agent velocity can be restated as:

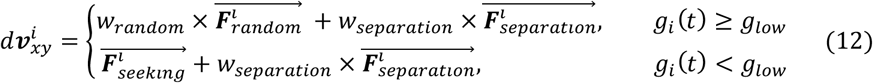

From here on, the simulation can be demonstrated and ready for some further epidemiological features.

### 3. Some epidemiological properties

When it comes to the simulated epidemics, agents can have an epidemiological state, be it susceptible, infectious, recovered, or dead, in a mutually exclusive manner. The state will be updated on each iteration according to some rules. As discussed previously, the coronavirus disease (COVID-19) is highly contagious from human to human. Via the droplets due to respiratory process, when people are close enough to talk or touch one another, or one human gets from droplets staying on surfaces as touched by hands as one-touch eyes, nose, or mouth. Physical distancing, sanitizing hands, and preventing touching faces are the campaign for people to resist the contagions. These aspects are the core thing to be simulated in our artificial societies.

Firstly we have a population with all susceptible agents, except the patient zero. The only susceptible agent can be infected due to her spatial interaction with other infectious agents, occurs with probability *p*_*infected*_ and only examined if the distance between the two agents is below a certain constant of *d*_*infection*_. After the *t*_*resolution*_-th iteration (distributed normally for all agents), infectious agent can be recovered with probability *p*_*recovery*_, otherwise, she is dead and removed from the artificial world. In return, recovered agents are still susceptible again with a particular probability *p*_*susceptible*_ after *t*_*susception*_-th iteration (also distributed normally).

### 3 Applying policies in the Artificial Society

Countries around the world manage the COVID-19 pandemic in various ways deliberating all aspects and social dimensions. The basic idea of strategy against COVID-19 in the absence of any pharmaceutical intervention is reducing the possibility of susceptible and infectious mix one another. This is brought by early ascertainment of cases or reduction of contact.

“Lockdown” is one of the most heard solutions since it was the practice in China during the first outbreak epidemic (Lau *et.al*., 2020). However, the basic idea for reducing the mixing of susceptible people with the infected one can be interpreted as applied to various enforced policies. The campaign of social distancing is one of them: people are given the understanding to be in the distance when around other people (Lewnward, *et. al*., 2020). This is including the order of not letting people be in gatherings in not more than a few. In most impacted regions and countries, the public spaces closures, *e.g*.: restaurant, recreational park, entertainment venues, market, school are forbidden to operate regularly. A late addition by the World Health Organization,s advice as a suggested intervention for the pandemic is universal face mask use. There are some rationales of the use of face masking as effective personal protective measures in the era of pandemic (Sunjaya & Jenkins, 2020).

Growing the epidemic *in silico* would be valuable in observing how some of those intervention strategies enforced in many countries. Four categorical types of interventions that can be observed in the artificial societies are social distancing encouragement, mask-wearing campaign, public spaces closures, and lockdown as presented in table 1. In its actuality, the four categorical types can be used as interventions in many places, countries, around the world in the existing combination as well.

**Table 1.** Type of interventions and the implementation in the model.

| Policies | Model implementation |
| --- | --- |
| Social distancing | In a social distancing scenario, all agents are expected to keep their distance from each other to reduce the disease transmissibility. This can be applied by adjusting the $d_{\text{separation}}$ value then see if the agents perform separation rule. If the separating distance among agents is greater than infection distance, no infection will occur in the probability of $(1-p_{\text{separate}})$, the $\mathbf{F}_{\text{separation}}$ is set to zero vector. |
| Mask-wearing | The mask-wearing scenario is simulated by multiplying the probability $p_{\text{infected}}$ with mask reducing factor $k = [0.1, 1]$. In the basic model, one can say that $k$ value is equal to 1 so that it will not affect the occurrence of infections. |
| Public spaces closures | This scenario is implemented by removing all the attraction points that previously exist in the basic model. The attraction force for each agent then automatically turns to zero vector. It's worth noting that this also affects the wellness index of agents where the distance between the agent and her nearest attraction point becomes infinity. |
| Lockdown | For the lockdown scenario, agents are redistributed to the initial vector in their “homes” then within the “home” uniform grids sized as $l_{\text{grid}} \times w_{\text{grid}}$ where each has centroid $\mathbf{r}_{\text{grid}}$. Once in her “home” position, the agent is attached to one of those grids in which the distance $d(\mathbf{r}, \mathbf{r}_{\text{grid}})$ is minimum. Whenever the agent wants to leave her grid, she will be pushed back by the wall so that she can't interact with other agents in another grid. In this scenario, all public spaces are omitted from the landscape. |

## 4 Discussions

In our artificial society, without the intervention at all, the dynamics of the epidemic rely only on the herd immunity formations within the population. However the aspects of the contagiousness of the COVID-19 would likely in demand of quite a long time to reach it since most fraction of the population should be infected to get there (Clemente-Suárez, *et. al*., 2020). There is also the risk of an unacceptable number of death tolls without any intervention.

Within the simulation, we check the popular “lockdown” intervention. The effective lockdown is impressively suppressed the number of infection rate. There are some social (and psychological) effects, nonetheless to the population in the period of locking down the population, the agent,s social and psychological index. This index can somehow represent the situation of social and economic aspects of the intervention since within the interval time people cannot do the social and activities at all effectively.

At the end of the day, locking-down the population needs accurate momentum along with social and economic aspects of individual and community life, especially when it comes to the timing of the reopening. The too-early reopening comes to the risk of the second wave. The world witnesses this situation in some countries of which reopening too soon of the measure. In order to suppress the second wave, another lock-down intervention should be enforced. In some places around the world, there are some cities and countries that need to do such measures.

When it comes to closing down the public spaces (in the simulation we omit the social attraction points) and encouraging the effective physical distancing measures to the population, the number infection rate is suppressed a little. But since the agents are still able to get along wandering within our landscape, the infections are still there. People are still getting infected and in the long run, the active cases are merely slowing down in a big period of time until the definitely possible herd-immunity comes along. This is shown in figure 5 (*left*).

**Figure 2.**
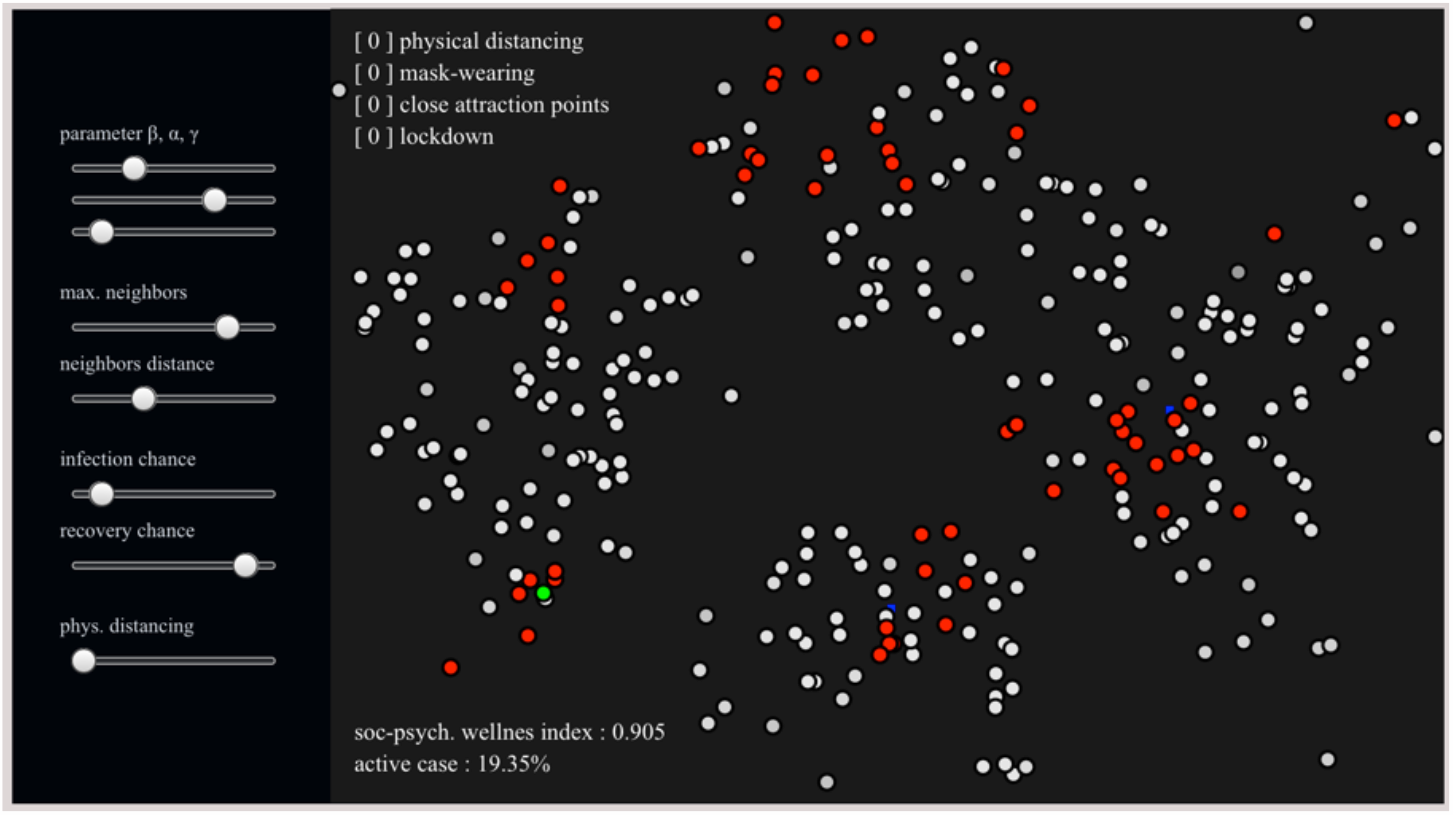
The screen of our artificial societies with the control panel for adjustments of the experiments we can deliver. The dots represent moving agents, while the red dots are infected, the green one represents the recovered agents, the white one is susceptible agents. The dark blue is the center of attraction in which agents fulfill their utmost social and psychological wellness.

**Figure 3.**
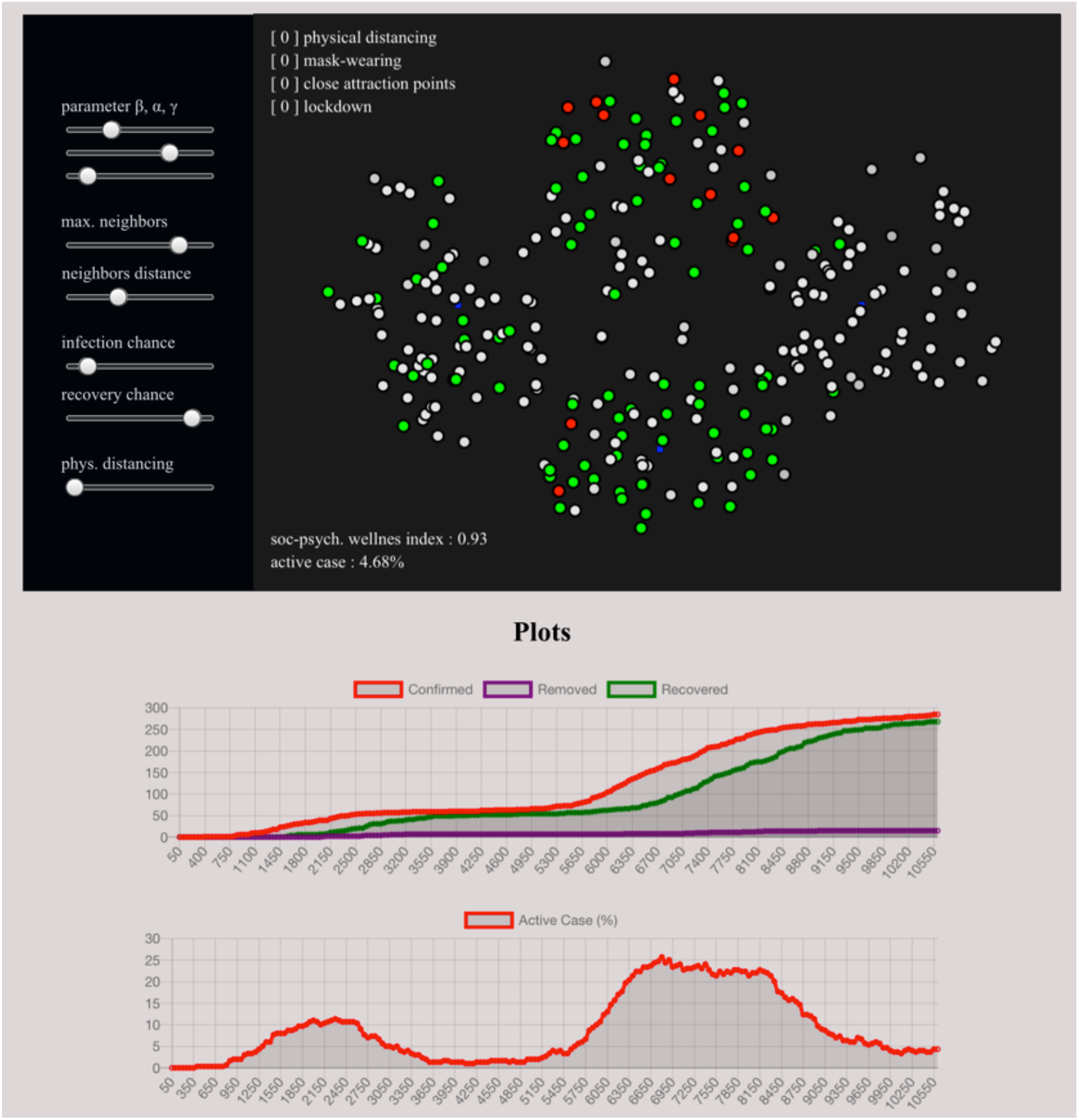
Pandemics in artificial societies, without intervention and the possibilities of the second wave.

**Figure 4.**
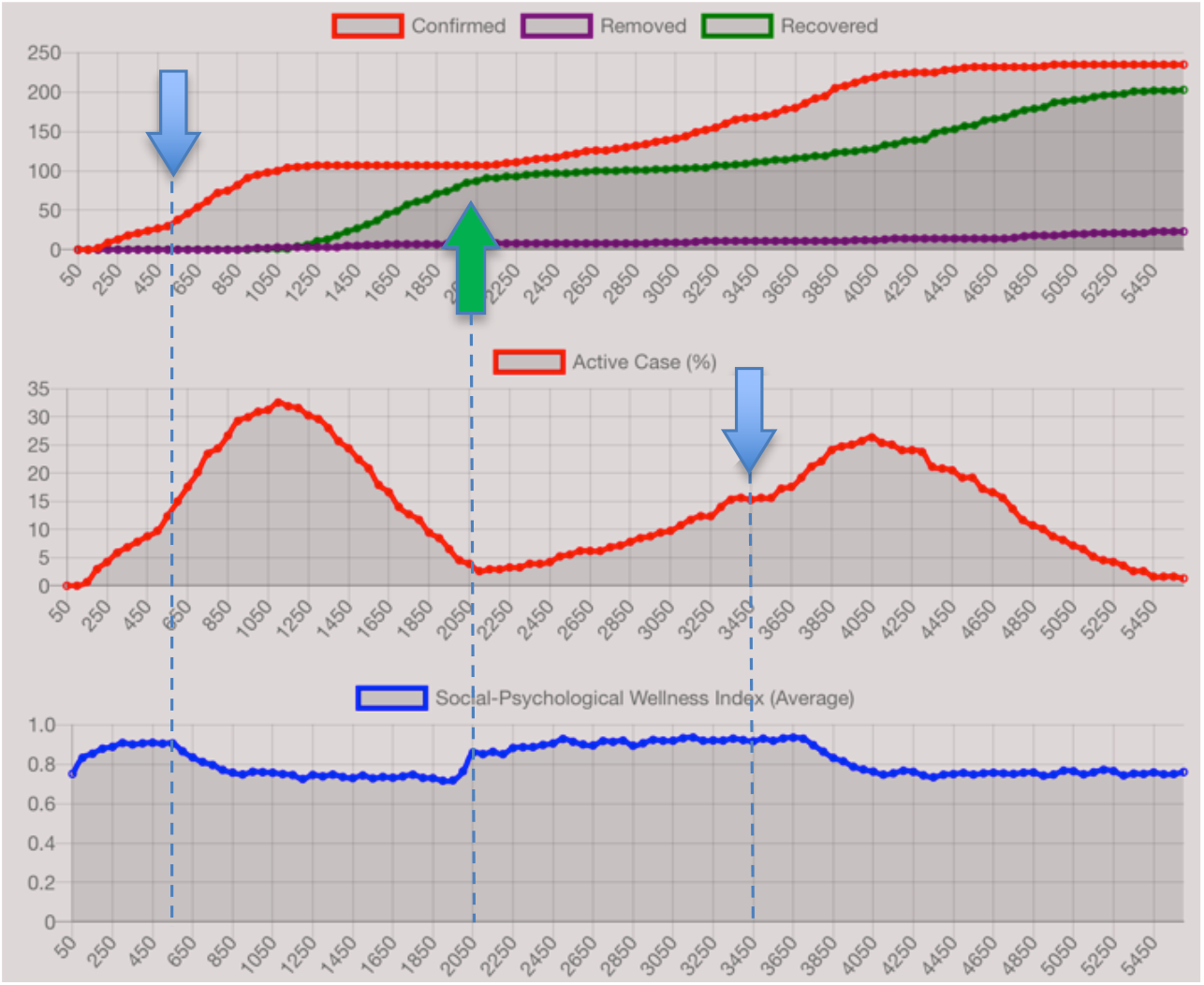
Effective locking-down the population (*green arrow*) suppressed the number of infection fast, with the risk of degrading of the average of the agent,s index of wellness. It is also simulated that wrong timing or too early re-opening the lockdown can severely emerge the second wave of the epidemic.

**Figure 5.**
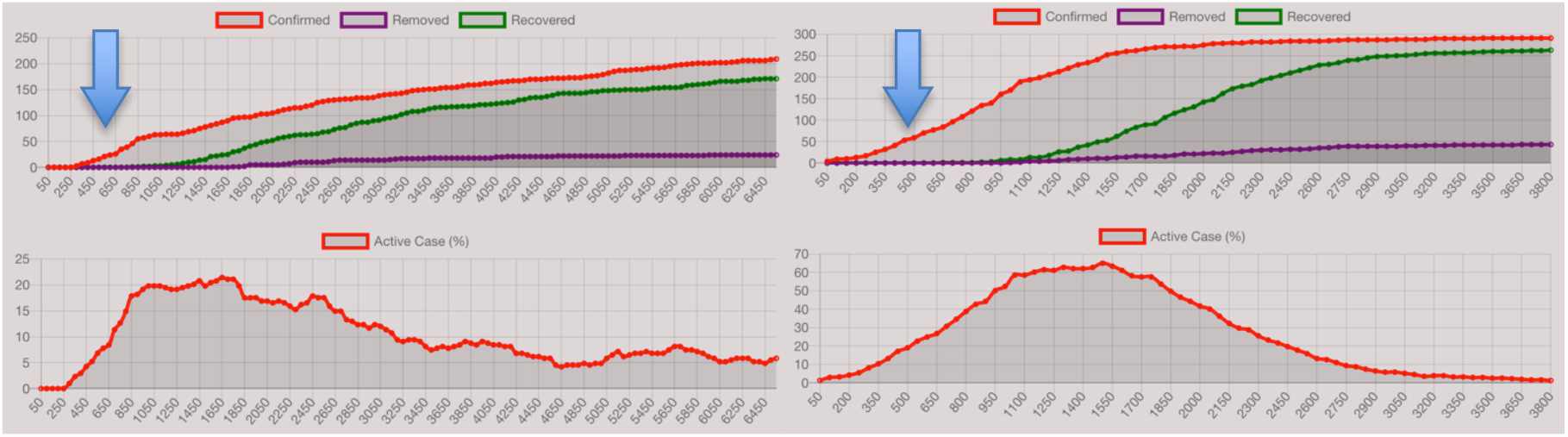
The intervention with social distancing and closing down all public spaces within the population (*left*) and the enforcement of mask usage within the population (*right*) are simulated.

As discussed earlier, the WHO later gives recommendations on the use (cloth) mask as personal protection in the era of a pandemic. As we simulated the usage of masks in our agent-based model, the slowing rate of infection does give effect even though it needs time to suppress the number of active cases. It is worth noting that the usage of masks in our simulation in figure 5 (*right*) is delivered exclusively without any other intervention.

The effectiveness of wearing masks with a combination of physical distancing encouragement and closure of public spaces for a period of time is simulated with the interesting results as shown in figure 6. The effectiveness of this combination, relative to the effective lockdown is the interval time for the cases eventually significantly decreased. However, since agents are still given the opportunity with their social and economic life, the things average social and psychological index is not as drop like the one with full effective lockdown intervention observed in the previous experiment. This may explain with the cases in South Korea, Taiwan, and some other eastern Asia, where people are encouraged strongly to wear masks even after the lockdown phase and the cases have been decreased (Lee & You, 2020).

**Figure 6.**
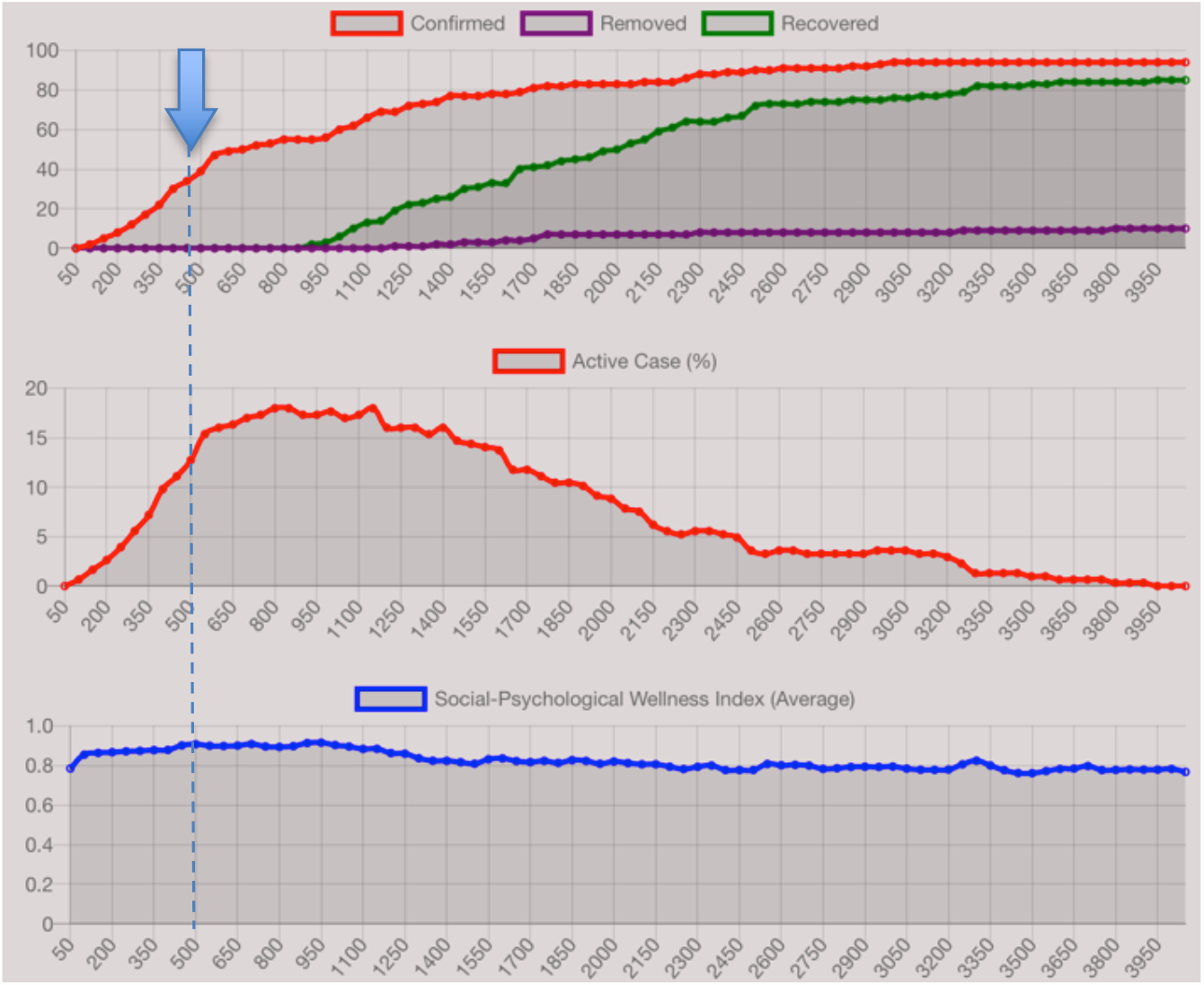
The intervention in the combination of public spaces closures, encouragement of physical distancing, and the wearing of masks.

Thus from our sets of experiments in the agent-based simulation, some tweaks of interventions due to the pandemic at the micro-level, the emerged macro-level is observed, including some emerged social aspects. There is no single solution of intervention when it comes to complex social systems, including due to the policy harnessing the pandemic. The computational simulation, whatsoever, is open for modifications and other changes due to many other aspects to be included in the intervention. Our simulation explains the varsities of governance and social and economic policies applied in different countries, regions, and areas.

## 5 Concluding Remarks

The agent-based model of the complex social system to observe some aspects of intervention due to the COVID-19 pandemic can give insights on the cause and emergence of intervention on trying to handle the pandemics in the absence of a vaccine. Some interventions potentially hurt the social and economy of the people while “flattening” the exponential rise of infections. On the other hand, no intervention can bring people with the risk of unacceptable death while naturally, the eco-social system adjusts itself for the collective immunity. On the other hand, the characteristics of the disease and virus are not the same either for different regions, areas, and countries.

Many aspects, not necessarily the social and economic one *per se*, should be put into account when it comes to policies. Computational simulation, by growing deliberately the structure of the social system into computation, provides the artificial societies in which many different observed aspects and characteristics of the disease can be applied (by tweaking variables in either micro and larger (macro) levels of description). Our simulation may explain and computationally demonstrated the various pathways of intervention delivered by the governments in different countries and regions.

## Data Availability

The computational simulation is provided online.

https://data.sains.club/simepi/

## ACKNOWLEDGMENTS

We thank colleagues in Bandung Fe Institute for discussions in the early draft of the manuscript.

## APPENDIX MODEL PARAMETERS

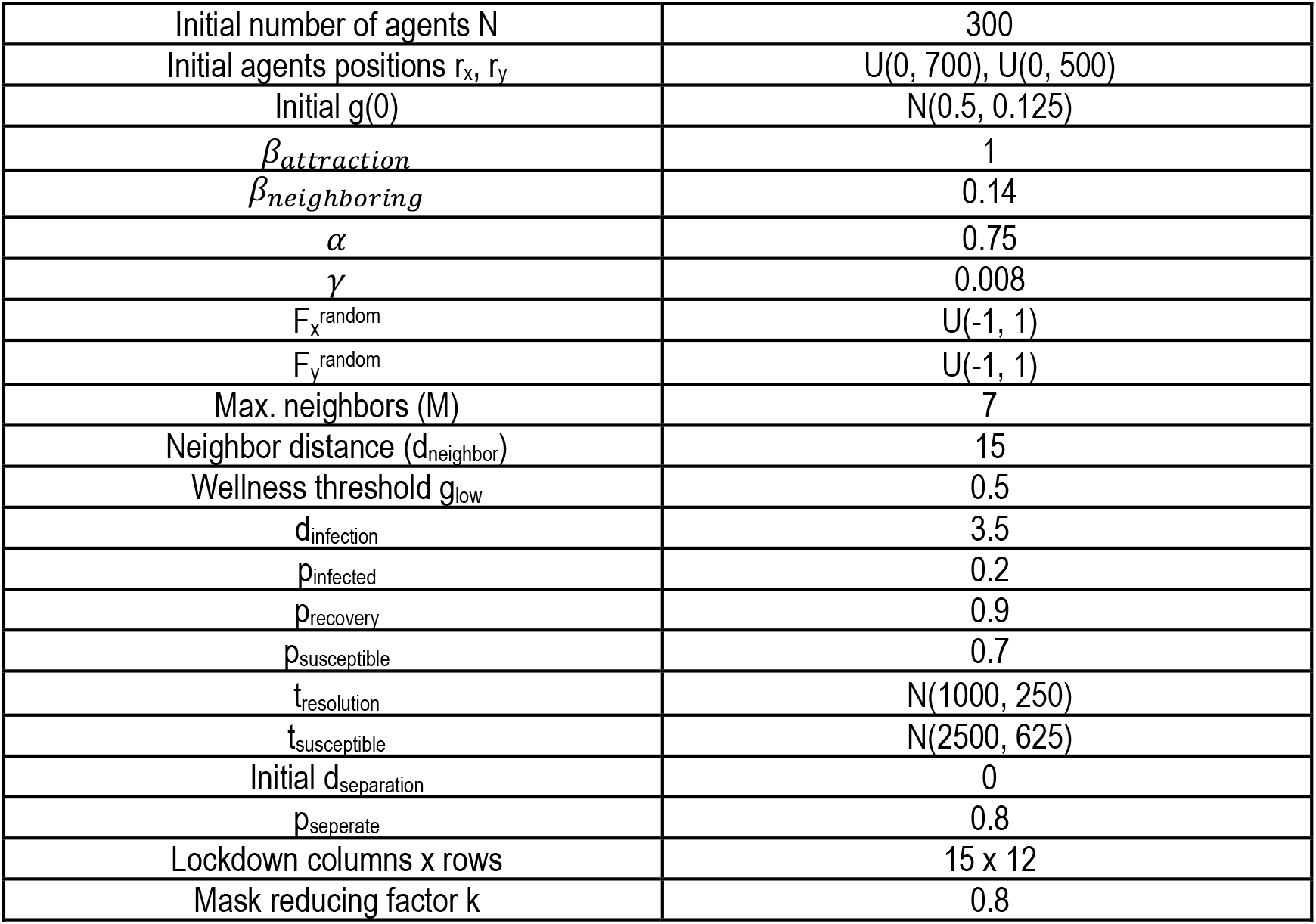

## Footnotes

1 https://www.nytimes.com/interactive/2020/world/asia/china-coronavirus-contain.html

